# Pairwise genetic meta-analyses between schizophrenia and substance dependence phenotypes reveals novel association signals

**DOI:** 10.1101/2021.09.12.21263471

**Authors:** Laura A. Greco, William R. Reay, Christopher V. Dayas, Murray J. Cairns

**Affiliations:** School of Biomedical Sciences and Pharmacy, The University of Newcastle, Callaghan, NSW, Australia; Centre for Brain and Mental Health Research, Hunter Medical Research Institute, New Lambton, NSW, Australia

**Author notes:** To whom correspondence should be addressed: Professor Murray Cairns, Medical Sciences Building, University Drive, Callaghan, NSW 2308, Australia.

## Abstract

Almost half of individuals diagnosed with schizophrenia also present with a substance use disorder, however, little is known about potential molecular mechanisms underlying this comorbidity. We used genetic analyses to enhance our understanding of the molecular overlap between these conditions. Our analyses revealed a positive genetic correlation between schizophrenia and the following dependence phenotypes: alcohol (*r*_*g*_ = 0.3685, *SE* = 0.0768, *P =* 1.61 × 10^−06^), cannabis use disorder (*r*_*g*_ = 0.309, *SE* = 0.0332, *P =* 1.19 × 10^−20^) and nicotine dependence (*r*_*g*_ = 0.1177, *SE* = 0.0436, *P =* 7.0 × 10-^03^), as well as lifetime cannabis use (*r*_*g*_ = 0.234, *SE* = 0.0298, *P =* 3.73 × 10^−^15) and drinks per week (*r*_*g*_ = 0.0688, *SE* = 0.0217, *P =* 1.5 × 10^−03^). We further constructed latent causal variable (LCV) models to test for partial genetic causality and found evidence for a potential causal relationship between alcohol dependence and schizophrenia (*GCP =* 0.6, *SE* = 0.22, *P =* 1.6 × 10^−03^). This putative causal effect with schizophrenia was not seen using a continuous phenotype of drinks consumed per week, suggesting that distinct molecular mechanisms underlying dependence are involved in the relationship between alcohol and schizophrenia. To localise the specific genetic overlap between schizophrenia and substance use disorders, we conducted a gene-based and gene-set pairwise meta-analysis between schizophrenia and each of the four individual substance dependence phenotypes in up to 790,806 individuals. These bivariate meta-analyses identified 44 associations not observed in the individual GWAS, including five shared genes that play a key role in early central nervous system development. These genes may play an important role in substance dependence in schizophrenia, and, as a result, could represent important targets for future treatment or early intervention, as comorbid substance dependence is associated with poor treatment adherence, greater chronicity and increased mortality.

## Introduction

Schizophrenia is a complex and debilitating psychiatric disorder with a diverse array of symptoms and comorbidities including substance dependence^1^. While the onset generally occurs during early adolescence, schizophrenia is highly heritable, which provides an opportunity to glean molecular insight into the composition of dysfunctional networks underlying its pathophysiology and phenotypic diversity, including associated comorbidities. Substance dependence (SD) is also a complex heritable trait with a polygenic architecture encompassing genetic variants across a multitude of genetic loci. Twin studies have provided insights into the magnitude of total SD heritability regardless of the substance^2^; alcohol (AD, 0.63), cannabis (CD, 0.78) nicotine (ND, 0.72), cocaine (CoD, 0.61) and opioids (OD, 0.61)^3^

While these substance use disorders arise independently of psychotic and affective disorders, they are frequently comorbid. A 2018 meta-analysis by Hunt *et al* of 123 articles between 1990 and 2017 (*n =* 165,811) found that the prevalence of any substance use disorder in schizophrenia was 41.7%, with 26.2% for cannabis, 24.3% for alcohol and 7.3% for stimulants^4^. Some population studies have reported even higher use, with 72% of people with schizophrenia being daily cigarette smokers^5^, and nearly half using cannabis regularly^6^. Although data on opioid use in schizophrenia is limited, it was found to be significantly higher than the general population^7^. Substance use and dependence is a significant complication for patients with schizophrenia and often impedes treatment options. For example, nicotine use increases haloperidol metabolism, such that smokers need higher doses compared to their non-smoking counterparts^8^. First-generation and short acting oral antipsychotics have also been shown to increase substance use and cravings, with alcohol use in patients worsening over time^9,10^.

Despite the high heritability and prevalence of SD in schizophrenia, Genome Wide Association studies (GWAS) for SD lag those for other major psychiatric disorders. In comparison to GWAS mega-analyses like schizophrenia and major depressive disorder, there is still the need for larger cohorts for SD phenotypes to boost discovery power, particularly for psychostimulants such as cocaine and amphetamines. Current GWAS on SD phenotypes have not revealed any clear novel biological pathways involved in these phenotypes, with limited loci found in SD, thus more work in this area is needed. In the current study we employed pair-wise meta-analysis GWAS to reveal genes potentially underpinning substance dependence in schizophrenia, that may not be apparent through investigation of the disorders individually. Understanding the molecular determinants of comorbid addiction in schizophrenia has the potential to identify new therapeutic targets, which may lead to improved clinical outcomes in those individuals with risk of these exacerbating conditions.

## Materials and methods

### GWAS

GWAS Summary statistics of European ancestry were obtained for Schizophrenia (*N =* 130,644) and Substance Dependence from the Psychiatric Genomics Consortium (PGC), International Cannabis Consortium (ICC) and Social Science Genetic Association Consortium (SSGAC). Substance dependence GWAS include Alcohol Dependence (unrelated genotyped individuals)^11^ (AD, *N =* 28,757), Nicotine Dependence^12^ (ND, *N =* 244,890 PMID: 31427789), Lifetime Cannabis Use excluding 23&me^13^, (LCU, *N =* 184,765), Cannabis Use Disorder (unrelated individuals)^14^ (CUD, *N =* 357,806), Opioid Dependence vs Unexposed Controls^15^ (OD, *N =* 28,709), Cigarettes per day^12^ (CPD, *N =* 90,143), and Drinks per week^16^ (DPW, *N =* 414,343).

### Genetic correlation and evidence of causation

Linkage disequilibrium score regression analysis (LDSC, v 1.0.1) was utilised to estimate genetic correlation between schizophrenia and each SD phenotype^17^. Common SNPs (MAF > 0.05) in the GWAS summary data were retained if they were available in the HapMap3 panel that excluded the MHC region and were not otherwise excluded by the ‘munge_sumstats.py’ script in the LDSC framework. Genetic correlation between two traits may be indicative of shared underlying biology but does not necessarily imply that the relationship is causal. To evaluate evidence for a causal relationship between SD phenotypes and schizophrenia, we constructed a latent causal variable model^18^ (LCV), as has been demonstrated elsewhere^18-21^. Briefly, the LCV model utilises the genome-wide SNP-trait association *Z* scores for two traits and the mixed fourth moments (cokurtosis) of the respective distributions to assess whether there is evidence for a causal effect of one trait on the other. As a result, the LCV framework can calculate a metric termed the posterior mean causality proportion (GCP), from which the sign can be used to infer a potential causal direction. In practice, GCP > 0 implies that trait one is partially genetically causal for the second trait, whilst GCP < 0 implies the reverse. Given GCP > 0, for example, marginal SNP-trait one effect sizes tend to be proportionally larger on trait two, but this is not observed in the opposite direction – hence, the direction of causal effect can be estimated as operating from trait one to trait two. We defined partial genetic causality using the recommended threshold of a significantly non-zero |GCP| > 0.6, as this was shown by O’Connor and Price in simulations to guard against false positives^18^. It should be noted that the posterior mean GCP is not an estimate of the magnitude of any potential causal relationship and should not be interpreted as such – rather it evaluates the strength of evidence for a putative causal relationship in either direction using genome-wide SNP effects.

### Gene-based Association Analysis

Gene-based association was performed on each disorder using MAGMA version 1.07 (https://ctg.cncr.nl/software/magma). The MAGMA gene-based method utilises *P*-values as input, whereby the test-statistic is a linear combination of SNP-wise *P*-values. In comparison to univariate GWAS, the burden of multiple testing correction is dramatically reduced in gene-based association analysis^22^. Gene-based association can also greatly boost power by signal aggregation across variants in the target regions when multiple causal variants influence the phenotype of interest^23^. The default gene-based test was used, a modified version of Brown’s method for combining *P* values such that test statistic inflation arising from SNP-wise dependency due to LD within genes can be suitably accounted for. The 1000 genomes phase 3 European reference panel is used as the LD reference for this purpose. Variants were mapped to 18297 autosomal protein-coding genes from NCBI hg19 genome-assembly. Genes that arise from the major histocompatibility complex (MHC, chr6:28477797–33448354) on Chromosome 6 were removed. Statistical inference for a significantly associated gene for each disorder was set as *P* < 2.7 × 10^−6^ to adjust for the number of genes tested via the Bonferroni method.

### Pairwise cross-disorder meta-analysis

The genic Z-score outputs from the gene-based association analysis for schizophrenia and each substance dependence phenotype (AD, CUD, ND, OD), which were probit transformation of the *P* values, were meta-analysed individually using MAGMA – that is, schizophrenia was meta-analysed with AD, then with CUD, and so on. A weighted Z-test was utilised for this purpose, which is based off the inverse normal (Stouffer’s) method, whereby the weight (wi) was set as the respective GWAS sample sizes^24^.

### Gene-set association analyses

Competitive gene-set analysis results were obtained using MAGMA. For this analysis, 14,969 hallmark, 2,921 canonical and 2598 regulatory miRNA target gene ontology gene-sets from the molecular signatures database (MsigDB, v7.4) were selected^25, 26^. A linear regression model is constructed by MAGMA wherein genic association (transformed to Z) is the outcome. Confounders that are adjusted for in this analysis include gene-size and genic-minor allele count. Gene-set association analysis was undertaken to find gene-sets where the common variant signal is enriched relative to all other genes considered. Genes that share biological or functional properties from a defined reference database are aggregated into sets that include molecular interactions, regulation, and products to determine pathways relevant to the phenotype of interest. Multiple testing correction for the gene-set analysis was performed using the Benjamini-Hochberg (BH) procedure, with FDR < 0.05 designated as a significant gene-set association^27^. For functional enrichment analysis of microRNA target gene-sets we used the web server g:Profiler.^28^

### Expression enrichment of implicated genes in the adult and developing brain

We performed gene-set enrichment analysis for discovery purposes on the genes that passed multiple testing correction for the schizophrenia and substance dependence (AD, CUD, ND, OD) pair-wise meta-analyses using the R-package ABAEnrichment (version 1.22.0)^29^. ABAEnrichment tests for expression enrichment in 414 adult brain regions and 26 brain regions at 5 different developmental stages from BrainSpan data set distributed by the Allen Brain Institute. The adult brain regions were sampled from a total of 6 donors, the 6 individuals were Caucasian (50%), Black or African American (33.3%), and Hispanic (16.6%). The 5 developmental stages included RNA-sequencing data from 42 individuals: prenatal (all stages), infant (0–2 years), child (3–11 years), adolescent (12–19 years), and adult (19-40 years). The 42 individuals were Caucasian (45%), Black or African-American (33%), Hispanic (10%), mixed (5%) and the other 5% had no ethnicity information available. The core function of the ABAEnrichment statistical analysis uses the program package FUNC^30^, which selects for hypergeometric testing for binary associated variable analysis. The multiple testing correction provided by FUNC was the family-wise error rate (FWER) method, which estimates the probability that at least one false positive category exists among those that are labelled significant. The web-based platform FUMA^31^ was used to further investigate the enrichment amongst different developmental periods.

## Results

### Evidence for partial genetic causality of alcohol dependence on schizophrenia

LD score regression (LDSC) analysis revealed significant genetic correlations between schizophrenia and several of the SD phenotypes after correcting for the number of tests performed. Specifically, schizophrenia was positively genetically correlated with AD *(r*_*g*_ = 0.3685, *SE =* 0.0768, *P =* 1.61 × 10^−06^), CUD (*r*_*g*_ = 0.3092, *SE* = 0.0332, *P =* 1.19 × 10^−20^) and ND (*r*_*g*_ = 0.1177, *SE =* 0.0436, *P =* 7.0 × 10^−03^). The substance use phenotypes drinks per week (*r*_*g*_ = 0.0688, *SE =* 0.0217, *P =* 1.5 × 10^−03^) and LCU (*r*_*g*_ = 0.234, *SE* = 0.0298, *P =* 3.73 × 10^−15^) also displayed significant non-zero schizophrenia correlation. We also found nominally positive genetic correlation between schizophrenia and OD (r_g_ = 0.1849, *SE =* 0.0754, *P =* 0.0142), and negative genetic correlation with cigarettes per day (r_g_ = -0.0388, *SE =* 0.0179, *P =* 0.0301), however, these two signals did not survive multiple testing correction. LCV models were then constructed for the significant traits for which their schizophrenia genetic correlation estimate passed multiple testing correction (AD, CUD, LCU, ND, and DPW) to investigate whether any of the observed genetic correlations between SD and schizophrenia may constitute a causal relationship (Supplementary Table 1). There was no evidence for partial genetic causality of CUD, LCU and ND on schizophrenia, but there was moderate evidence that AD was partially genetically causal for schizophrenia (*GCP =* 0.60, *SE =* 0.22, *P* = 0.001), whilst DPW did not show any evidence for a causal relationship with schizophrenia like alcohol dependence. Interestingly, AD and DPW showed genetic correlation (*r*_*g*_ = 0.6, *SE =* 0.099, *P =* 1.31 × 10^−09^) but there was no evidence for partial genetic causality of DPW on AD (*GCP =* 0.27, *SE =* 0.45, *P* = 0.55), suggesting that the underlying mechanisms driving AD may not present in DPW observed in a population sample, although this requires further investigation.

**Fig 1.**
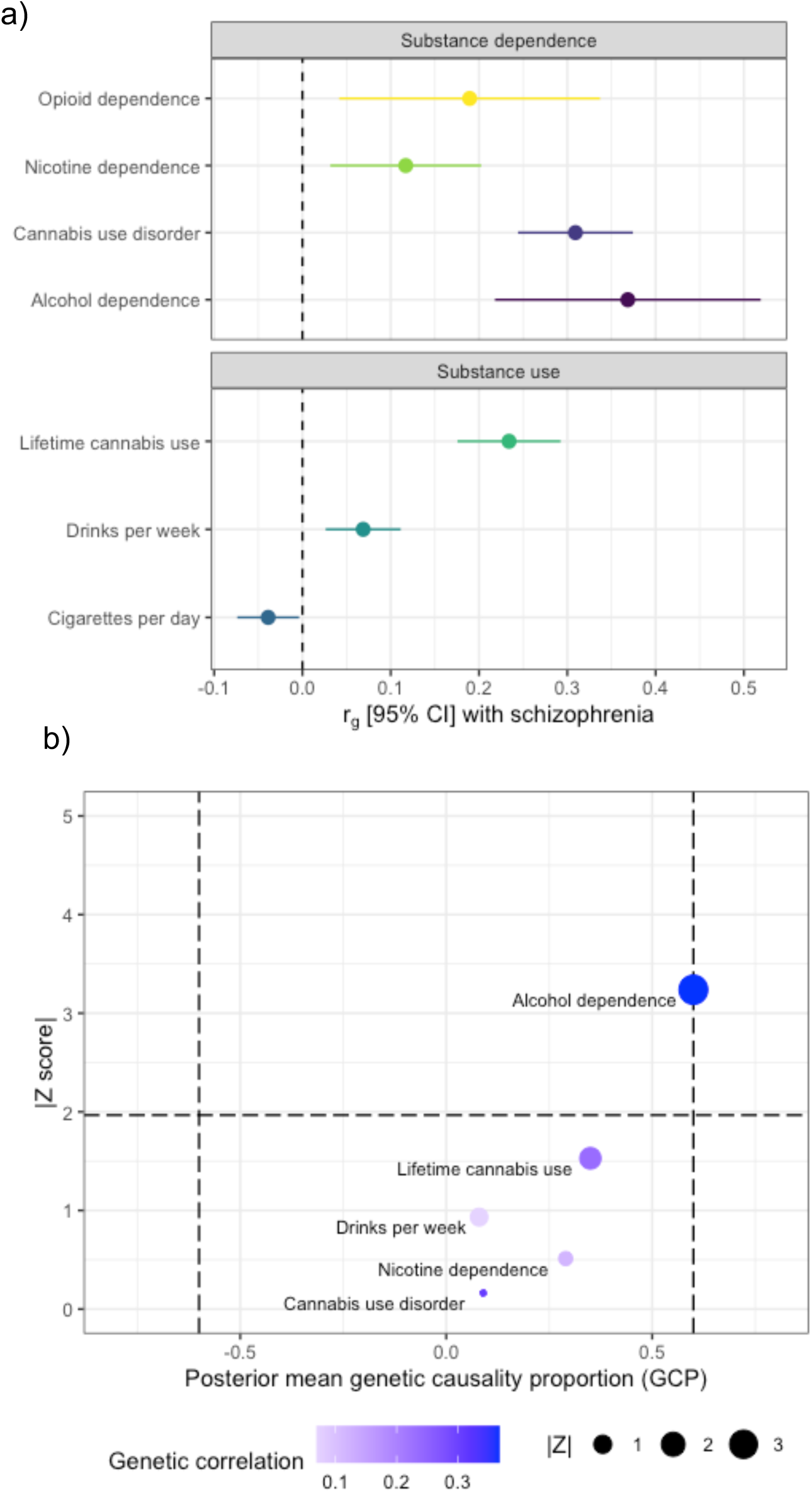
**a)** Genetic correlation forest plot between schizophrenia and SD (AD, CUD, ND, and OD), rg calculated using linkage disequilibrium (LD) score. **b)** Latent causal variable (LCV) model to test for partial causality with GCP estimates of SD on schizophrenia.

### Gene based multivariate association reveals 44 novel signals for substance dependence and schizophrenia

Using aggregated gene-level association (MAGMA) on the individual traits, we observed that 534 genes were associated with schizophrenia alone after Bonferroni correction (P < 2.7 × 10^−6^, Supplementary Table 2). For cannabis use disorder, 2 genes were significantly associated including, *PDE4B* (*P =* 2.09 × 10^−06^) and *FOXP2* (*P =* 9.30 × 10^−07^, Supplementary Table 3), and one gene for nicotine dependence – *ARHGAP22* (*P =* 2.42 × 10^−6^, Supplementary Table 4). While no genes were associated with AD and OD after multiple testing correction, 9 genes were significantly associated with life-time cannabis use including, *LRRTM4* (*P =* 4.51 × 10^−07^) *CADM2* (*P =* 1.14 × 10^−13^) *AS3MT* (*P =* 9.43 × 10^−07^) *NCAM1* (*P =* 1.44 × 10^−09^) *ATXN2L* (*P =* 1.85 × 10^−08^) *TUFM* (*P =* 5.72 × 10^−08^) *SH2B1* (*P =* 3.15 × 10^−08^) *ATP2A1* (*P =* 1.68 × 10^−08^) *RABEP2* (*P =* 1.64 × 10^−06^) and *SRR* (*P =* 1.61 × 10^−06^) (Supplementary Table 5) with none of these genes shared with CUD.

The genetic architecture shared between schizophrenia and substance dependence was further investigated using genic pairwise meta-analysis (MAGMA) to identify potentially pleiotropic signals that do not reach conventional significance thresholds in the individual GWAS. The number of genes which survived Bonferroni correction in each of the meta-analyses was as follows: 444 genes for schizophrenia meta-analyses with OD. 437 genes for schizophrenia meta-analysed with AD, 94 genes for schizophrenia meta-analysed with CUD, and 99 for schizophrenia meta-analysed with ND. The top hit for schizophrenia, *HIST1H4* (*P* = 2.86 ×10^−38^), was also the most significant for the schizophrenia meta-analysis with AD (*P* = 2.70 ×10^−31^), and OD (*P* = 4.50 ×10^−36^). Whereas the most significant gene observed in the CUD and schizophrenia meta-analysis was *ST3GAL3* (*P* = 9.68 ×10^−16^) and in the ND and schizophrenia meta-analysis the gene *ZFYVE21* (*P* = 1.11 ×10^−14^). We then restricted these genes to those which were also at least nominally significant (*P*<0.05) in the individual GWAS for schizophrenia and SD but did not survive multiple testing correction for either of the respective univariable GWAS (schizophrenia+AD = 16, schizophrenia+CUD = 15, schizophrenia+ND = 5, schizophrenia+OD = 13, Supplementary Table 6-9). These genes are likely more biological salient, as many of the other which survived correction in the meta-analyses that did not reach nominal significance in the respective SD GWAS were driven purely by schizophrenia given its greater discovery power. Interestingly, five of these genes were found in more than one bivariate meta-analysis; specifically, (1) *TRAF3IP2* and (2) *MED19* in the schizophrenia+AD, schizophrenia+CUD, and schizophrenia+ND meta-analyses; (3) and BDNF, (4) *FUT2* and (5) *IZUMO1* for both the schizophrenia+AD and schizophrenia+CUD meta-analysis.

**Table 1.** The top ten significantly associated genes observed in the schizophrenia and substance dependence (AD, CUD, ND, and OD) MAGMA meta-analyses that were nominally significant (P<0.05) in the individual analysis but did not pass multiple testing corrections.

| GENE SYMBOL | $P_{SZ}$ | $P_{SD}$ | $P_{META}$ |
| --- | --- | --- | --- |
| MED27 | $6.47 \times 10^{-5}$ | $1.425 \times 10^{-5}$ | $8.64 \times 10^{-9}$ |
| BDNF | $4.22 \times 10^{-6}$ | AD, $1.08 \times 10^{-2}$<br>CUD, $1.39 \times 10^{-4}$ | AD, $2.75 \times 10^{-7}$<br>CUD, $3.42 \times 10^{-8}$ |
| IZUMO1 | $2.29 \times 10^{-5}$ | AD, $6.52 \times 10^{-3}$<br>CUD, $1.13 \times 10^{-4}$ | AD, $1.05 \times 10^{-6}$<br>CUD, $7.67 \times 10^{-8}$ |
| HYAL3 | $2.06 \times 10^{-5}$ | $1.47 \times 10^{-4}$ | $9.83 \times 10^{-8}$ |
| HMGN4 | $3.65 \times 10^{-4}$ | $2.63 \times 10^{-5}$ | $1.00 \times 10^{-7}$ |
| EP300 | $1.18 \times 10^{-5}$ | $9.72 \times 10^{-4}$ | $1.36 \times 10^{-7}$ |
| SPECC1 | $8.28 \times 10^{-6}$ | $1.99 \times 10^{-3}$ | $1.51 \times 10^{-7}$ |
| PPP2R2A | $2.95 \times 10^{-6}$ | $9.82 \times 10^{-3}$ | $1.78 \times 10^{-7}$ |
| MED19 | $1.16 \times 10^{-5}$ | AD, $2.82 \times 10^{-2}$<br>CUD, $3.53 \times 10^{-4}$ | AD, $1.74 \times 10^{-6}$<br>CUD, $2.01 \times 10^{-7}$ |
| FUT2 | $3.37 \times 10^{-6}$ | $1.02 \times 10^{-2}$ | $2.10 \times 10^{-7}$ |

**Fig 2.**
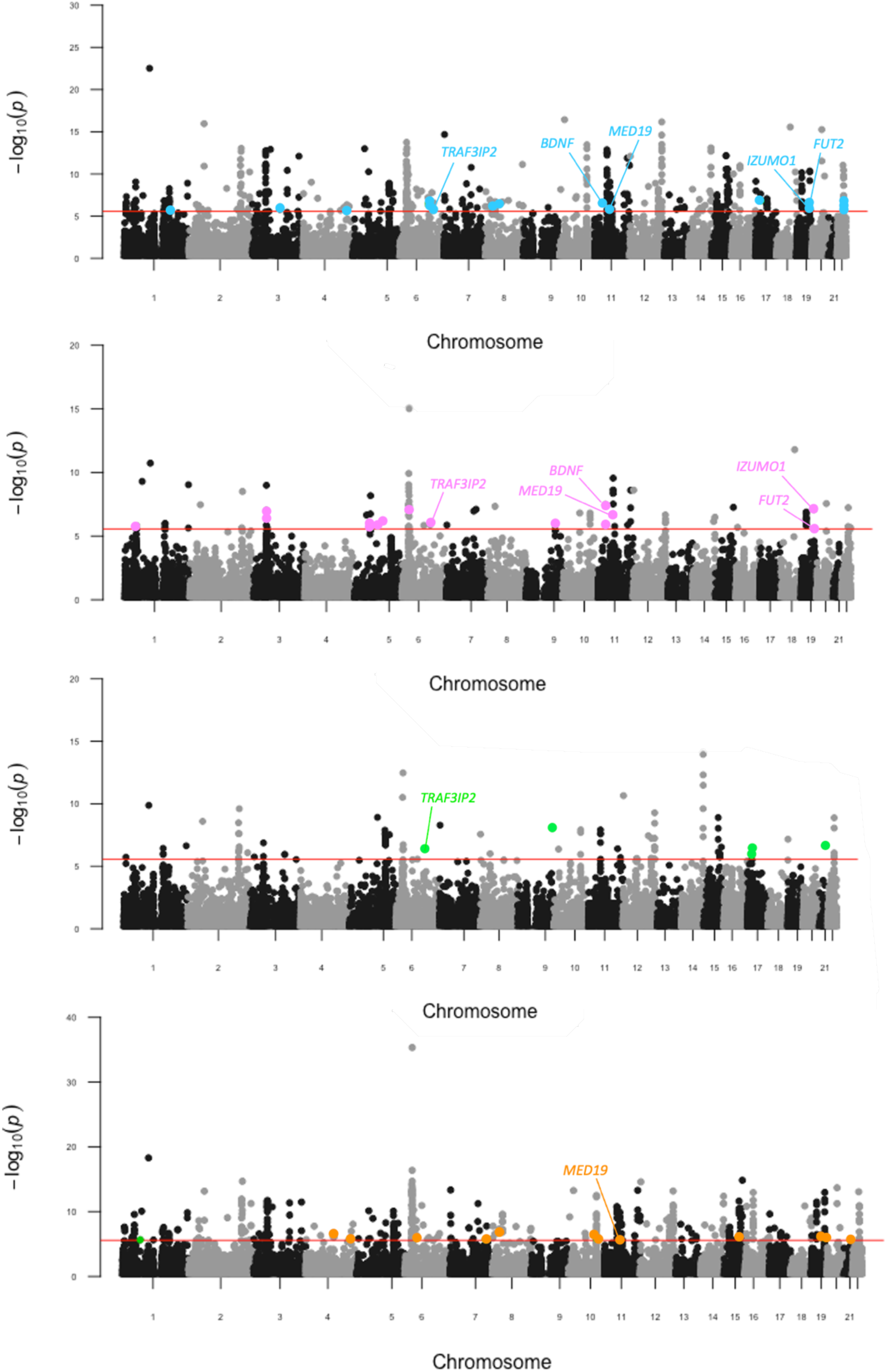
Pairwise genic meta-analyses of schizophrenia and SD. Manhattan plot for each meta-analysis which displays the −log10-transformed P value for association for genes which were tagged by at least one SNP in the respective GWAS. The red line represents the Bonferroni threshold for multiple-testing correction (P < 2.7 × 10−6). Genes highlighted on each plot were not Bonferroni significant in the individual GWAS, with overlapping genes across meta-analyses labelled. **a**. SZand AD, **b**. SZ and CUD **c**. SZ and ND. d. SZ and OD.

### Bivariate gene-set association meta-analysis uncovers novel biological systems involved in schizophrenia and substance use disorders

We investigated the involvement of 14,969 gene sets (MsigDB v7.4) in four categories including Biological Process (BP), Cellular Component (CC), Molecular Function (MF) and Human Phenotype Ontology (HPO), respectively. For the individual schizophrenia GWAS, 84 gene sets remained statistically significant after multiple testing correction (FDR <0.05). No gene-sets passed false discovery rate (FDR) correction for the individual substance dependence phenotypes.

In the pair-wise meta-analysis, 73 gene sets were statistically significant (FDR <0.05), for schizophrenia meta-analysed with AD, with 10 gene-sets not previously observed in the individual schizophrenia GWAS (Supplementary Table 10), including *long term synaptic potentiation* (n_*genes*_ = 81, *P =* 5.71 × 10^−05^, *FDR =* 0.02), *exocytic vesicle* (n_*genes*_ = 196, *P =* 1.23 × 10^−04^, *FDR =* 0.03), *paroxysmal ventricular tachycardia* (n_*genes*_ = 23, *P =* 1.70 × 10^−04^, *FDR =* 0.03), *peptidyl serine dephosphorylation* (n_*genes*_ = 19, *P =* 1.75 × 10^−04^, *FDR =* 0.04), *hypoplasia of the olfactory bulb* (n_*genes*_ = 4, *P =* 2.12 × 10^−04^, *FDR =* 0.04), *regulation of heart contraction* (n_*genes*_ = 221, *P =* 2.32 × 10^−04^, *FDR =* 0.04) and *regulation of peptidyl serine dephosphorylation* (n_*genes*_ = 5, *P =* 2.32 × 10^−04^, *FDR =* 0.04). The schizophrenia and CUD meta-analysis yielded 33 statistically significant sets, with 12 not observed in the individual SCZ GWAS (Supplementary Table 11). Some of these novel gene-sets of interest were; *abnormal social behaviour* (n_*genes*_ = 140, *P =* 7.46 × 10^−07^, *FDR =* 0.002), *aggressive behaviour* (n_*genes*_ = 165, *P =* 1.75 × 10^−05^, *FDR =* 0.018), *hypoplasia of the olfactory bulb* (n_*genes*_ = 4, *P =* 7.24 × 10^−06^, *FDR =* 0.01), and *obsessive compulsive behaviour* (n_*genes*_ = 90, *P =* 6.66 × 10^−05^, *FDR =* 0.04). There were 60 gene-sets passed FDR correction in the schizophrenia meta-analysis with OD (Supplementary Table 12), with 9 not previously seen in individual SCZ GWAS, including *spherical high density lipoprotein particle* (n_*genes*_ = 8, *P =* 6.84 × 10^−05^, *FDR =* 0.02), *histone deacetylase complex* (n_*genes*_ = 70, *P =* 9.25 × 10^−05^, *FDR =* 0.03) and *aggressive behaviour* (n_*genes*_ = 165, *P =* 1.68 × 10^−05^, *FDR =* 0.04). Finally, schizophrenia meta-analysed with nicotine dependence yielded the fewest significant gene-sets (31 gene-sets, Supplementary Table 13), however, 10 sets were still novel relative to what was observed in each individual GWAS, such as; *neurotrophin receptor binding* (n_*genes*_ = 11, *P =* 2.85 × 10^−05^, *FDR =* 0.02), *glutamatergic synapse* (n_*genes*_ = 284, *P =* 4.00 × 10^−05^, *FDR =* 0.03), *long term synaptic potentiation* (n_*genes*_ = 81, *P =* 4.69 × 10^−05^, *FDR =* 0.03) and *type I pneumocyte differentiation* (n_*genes*_ = 5, *P =* 7.70 × 10^−05^, *FDR =* 0.03). Interestingly, the human behaviour ontology set *abnormal aggressive impulsive* or *violent behaviour* and *aggressive behaviour* was common amongst schizophrenia meta-analysed with CUD, ND or OD, whilst *long term synaptic potentiation (LTP)* was also common between schizophrenia and AD and ND.

We then considered the association of 2,598 microRNA (miRNA) regulatory target prediction gene-sets with each individual GWAS, followed by the bivariate meta-analyses (Supplementary Tables 14-18). There were 239 miRNA that passed FDR correction for the individual schizophrenia GWAS, no miRNA regulator target gene-sets passed FDR correction for any of the individual substance dependence phenotypes. Notably, each of the meta-analyses revealed a total of 17 microRNA regulatory target gene-sets, not seen in the individual phenotypes, including six found in more than one bivariate meta-analysis. One such interesting example was the predicted target genes of miR-495, a microRNA that is highly enriched in the nucleus accumbens and has been shown to play a role in addiction related behaviours (PMID: 28044061). MiR-495 survived correction in both schizophrenia and AD meta-analysis (n_*genes*_ = 231, *P =* 2.43 × 10^−03^, *FDR =* 0.03) and the schizophrenia and ND model (n_*genes*_ = 231, *P =* 5.88 × 10^−05^, *FDR =* 0.01) but was only nominally significant in the individual schizophrenia GWAS (P = 0.007), supporting how this meta-analysis approach can increase discovery power.

The miRNA miR-137 is one of the most well studied genes implicated by schizophrenia GWAS, the regulatory targets of miR-137 (n_*genes*_ = 487) were significantly enriched with schizophrenia associated variation considering the individual schizophrenia GWAS (*P =* 4.54 × 10^−05^, *FDR =* 0.004). However, meta-analysis of schizophrenia with each of the substance dependence phenotypes did not increase this association, suggesting the signal was more localised to schizophrenia upon considering the currently available GWAS. We hypothesised that the biology associated with targets of miR-137 may be shared with that of the 17 novel miRNA target sets implicated by the meta-analysis. To test this, we evaluated which biological pathways and other ontological gene-sets the targets of miR-137 were overrepresented in relative to the 17 novel miRNA target sets uncovered. We found that there were several neuronal related processes enriched amongst the targets of miR-137 and at least one of the 17 miRNAs’, such as *neurogenesis, nervous system development*, and *glutamatergic synapse* and several implicated in metabolic processes.

### Early developmental periods (0-2yrs) revealed as crucial time-points by expression enrichment of developing brain

The genes that passed multiple testing correction in the pair-wise meta-analyses for schizophrenia and AD (*n*_*genes*_ = 437), schizophrenia and CUD (*n*_*genes*_ = 94), schizophrenia and ND (*n*_*genes*_ = 99), and schizophrenia and OD (*n*_*genes*_ = 444), were annotated to 414 regions in the adult brain and 26 regions across five developmental stages. The brain regions with genes in the top 10% of expression levels (cut-off >90%, FWER <0.05) were selected to provide valuable insight into the biological role these risk genes may play in brain functionalities and highlight developmental periods that incur vulnerability.

In age category 1 (prenatal - all stages), only one region, the primary motor cortex (area M1, area 4), was significant in the individual schizophrenia GWAS, no genes were over-represented in any of the individual substance dependence GWAS or the schizophrenia and substance dependence meta-analyses. In age category two (infant, 0–2 years), the primary visual cortex (striate cortex, area V1/17) and anterior (rostral) cingulate (medial prefrontal) cortex was over-represented in the individual schizophrenia GWAS. In schizophrenia meta-analysed with each of AD, OD and ND, the inferolateral temporal cortex (area TEv, area 20) was an over-represented region, as was the posteroventral (inferior) parietal cortex in both schizophrenia and OD and schizophrenia and ND. For the child age category (3–11 years) and for adolescent (12–19 years), no structures were overrepresented in the individual schizophrenia GWAS or the pair-wise meta-analysis for AD, CUD, ND, and OD.

In the adult brain, age category (19 – 40), no structures were overrepresented in the individual schizophrenia and substance dependence GWAS or the pair-wise meta-analysis for AD, CUD, ND, and OD. These results suggest that early brain developmental periods may play the largest role in genetic vulnerability to substance dependence in schizophrenia. Functional mapping of differentially expressed genes (DEG) to 29 different ages of brain samples using FUMA, showed significant down-regulation (*P*_Bonferroni_ <0.05) of the schizophrenia and SD meta-analysis gene-sets between 8 post-conception weeks (PCW) and 2 years old.

## Discussion

In this study we used genetic approaches to explore common genes between schizophrenia and substance dependence. We found strong evidence of genetic correlation between schizophrenia and these dependence phenotypes, whilst there was further evidence of a causal relationship between alcohol dependence and schizophrenia. Interestingly, there was no causal relationship between the consumption of alcohol (drinks per week) and schizophrenia, or between AD and DPW. This was consistent with the trans-ancestral GWAS of alcohol dependence^11^, which suggested that there is a distinction in the underlying molecular mechanisms driving pathological and non-pathological behaviours for substance use and dependence. It is well known that psychotic symptoms can occur in several clinical conditions related to alcohol such as intoxication, withdrawal, alcohol-induced psychotic disorder, and delirium ^32^. A study on 18,478 Finnish inpatients found alcohol-induced psychosis was the most common type of substance-induced psychotic disorders (SIPD)^33^ with the eight-year cumulative risk at 5.0% (95% CI, 4.6%-5.5%). A separate Swedish study that followed 7,606 individuals for 84 months between 1995 and 2015 to study the progression of schizophrenia from SIPD found that for alcohol the risk was of SIPD was 4.7%^34^. Interestingly, the 20-year conversion rate for patients with alcohol induced psychosis to schizophrenia was much higher, at 22.1%^35^ (95% CI=17.6−27.5). The putative causal relationship of AD on schizophrenia warrants further epidemiological and biological interrogation. There are also some important limitations to the use of LCV models – specifically, they are bivariate in nature, and thus, cannot model the effect of other plausible mediators or confounders, whilst the posterior mean GCP estimate is also not a causal estimate that could be afforded by approaches like Mendelian randomisation^18^. However, the use of Mendelian randomisation with a binary exposure like AD can be challenging^36^, particularly as only a handful of genome-wide significant SNPs have been identified that could be suitable instrumental variables.

Strikingly, we also observed 44 genes associated with substance dependence in schizophrenia that were not seen in the individual GWAS. Five of these genes (*TRAF3IP2, MED19, BDNF, FUT2* and *IZUMO1)* were common hits in more than one of the paired disorders. *TRAF3IP2*, encodes nuclear factor-kappa-B (NF-κB) activator 1 (Act1), an IL-17 receptor adaptor protein^37^. *TRAF3IP2* plays a critical role in the activation of multiple pro-inflammatory signalling pathways^38^, particularly IL-17. IL-17 is a negative regulator of adult hippocampal neurogenesis, with the absence of IL-17 shown to significantly improve neurogenesis and enhance synaptic function^39^. Prenatal IL-17 expression has been shown to influence neurodevelopment because of its role in cell differentiation, signalling and survival^39^. Mediator complex subunit 19 (*MED19*) is a physical and functional target of RE1 silencing transcription factor (REST), with combined depletion of MED19/MED26 shown to result in de-repression of REST targets in vivo^40^. REST is the master transcription factor of neuron-specific genes, particularly during postnatal brain development^41^ and has been shown to modulate μ-opioid receptor (MOR) gene expression^42^. The Mu opioid receptor (MOR) is a key modulator of the dopaminergic system, with the rewarding properties of opioids and non-opioid drugs shown in Oprm1−/− mice to be reduced or eliminated^43^.

*MED19* is also a mediator of peroxisome proliferator-activated receptor gamma (PPARγ) transcriptional activity^44^ which is essential for adipogenesis and glucose uptake and storage^45^. Interestingly, in the schizophrenia and OD gene-set analysis, the gene-set *spherical high density lipoprotein particle* was enriched. In humans, activation of PPARγ is generally associated with an increase in plasma HDL-cholesterol^46^. Adipose tissue is an endocrine gland, which secretes leptin and adiponectin, cytokines with pro- and anti-inflammatory properties, respectively^47^. The dysregulation of adipokine levels has been associated with schizophrenia and other related disorders^48^, with Olanzapine, clozapine, and quetiapine shown to elevate the pro-inflammatory cytokine, leptin^49^. Cocaine- and amphetamine-regulated transcript (CART) production and action is also modulated by leptin^50^. Moreover, there is evidence that schizophrenia genetic risk amongst insulin and glycaemic related pathways could be a target of therapeutic intervention^51, 52^.

Brain-derived neurotrophic factor (BDNF) has long been linked to neurodegenerative diseases and psychiatric disorders such as substance dependence^53^. BDNF plays an important role in the regulation of synaptic strength and plasticity in the brain^54^, glucose metabolism and the regulation of mammalian food intake via signalling in hypothalamic circuits^55, 56^. Hyperphagic obesity has been shown to develop in humans heterozygous for BDNF^57, 58^. Notably, we also detected a novel, Bonferroni significant association with *BDNF* upon meta-analysis of schizophrenia with alcohol dependence and cannabis use disorder. *BDNF* has not been previously detected in hypothesis-free association studies of these phenotypes. In the gene-set analyses, *neurotrophin receptor binding* was also a novel ontological pathway significant for schizophrenia and ND. Additionally, several of the novel 44 genes have also been found to play important roles in metabolism. In mouse studies, protein tyrosine kinase 2 beta (PTK2B) was found to play a critical role in the differentiation of beige adipocytes^59^, CD47 -/- knockout resulted in resistance to insulin desensitisation, glucose intolerance and diet-associated weight gain^60^. *Spherical high density lipoprotein particle* was also a significant pathway in the Schizophrenia and OD meta-analysis.

The ontologies enriched for the 17 microRNA regulatory target gene-sets not seen in the individual phenotypes, that were significant in the schizophrenia and substance dependence analyses also revealed some biologically salient insights. For instance, miR-5580 target genes were enriched in *insulin receptor signalling pathway* and miR-7856 target genes were enriched in several pathways implicated in the regulation of glucose metabolism. miR-137 target gene-sets were also enriched in several metabolic pathways including r*egulation of cellular response to insulin stimulus*. Literature has shown that microRNAs may regulate gene networks involved in disorders like schizophrenia^61^. miR-495 also directly targets the 3’UTR of BDNF, with overexpression of miR-495 found to suppress cocaine self-administration in mice.^62^

Despite the significant disease comorbidity in schizophrenia and substance dependence, previous GWAS for these disorders failed to reveal large overlap between genome-wide significant hits. We demonstrate that the increase in power afforded through our pair-wise meta-analysis approach was able to identify shared genetic signals, including new genes and biological pathways relevant to both the neurobiology of addiction and psychosis. These novel genes and biological systems should be used to future analyses to refine whether causal variation is mapped to these genes, and whether any of these signals could warrant therapeutic intervention.

## Data Availability

All data used in this study were extracted from public GWAS summary statistics.

## Acknowledgements

This study was supported by NHMRC project grants (1147644 and 1188493). L.A.G is supported by a University of Newcastle Research Scholarship (UNRS). W.R.R is supported by an Australian Government Research Training Program Stipend. M.J.C. is supported by an NHMRC Senior Research Fellowship (1121474), and a University of Newcastle College of Health Medicine and Wellbeing, Gladys M Brawn Senior Fellowship.

## Data and code availability

All data used in this study were extracted from public GWAS summary statistics.

